# Integrative Bioinformatics Analysis Provides Insight into the Molecular Mechanisms of 2019-nCoV

**DOI:** 10.1101/2020.02.03.20020206

**Authors:** Xiang He, Lei Zhang, Qin Ran, Junyi Wang, Anying Xiong, Dehong Wu, Feng Chen, Guoping Li

## Abstract

The 2019-nCoV is reported to share the same entry (ACE2) as SARS-CoV according to the updated findings. Analyzing the distribution and expression level of the route of coronavirus may help reveal underlying mechanisms of viral susceptibility and post-infection modulation. In this study, we found that the expression of ACE2 in healthy populations and patients with underlying diseases was not significantly different, suggesting relatively similar susceptibility. Besides, based on the expression of ACE2 in smoking individuals, we inferred that long-term smoking might be a risk factor for 2019-nCoV. Analyzing the ACE2 in SARS-CoV infected cells suggested that ACE2 was more than just a receptor but also participated in post-infection regulation, including immune response, cytokine secretion, and viral genome replication. Moreover, we also constructed Protein-protein interaction (PPI) networks and identified hub genes in viral activity and cytokine secretion. Our findings may explain the clinical symptoms so far and help clinicians and researchers understand the pathogenesis and design therapeutic strategies for 2019-nCoV.

## Introduction

In early December 2019, several cases of pneumonia with unknown causes were reported in Wuhan, Hubei, China(1). Later, a novel coronavirus, 2019-nCoV was identified based on sequencing of the patient respiratory tract samples. As of February 1, 2020, more than 10,000 confirmed patients and more than 15,000 suspected cases were reported in China, with additional patients being identified in a rapidly growing number internationally. Analyzing of the genome of 2019-nCoV showed that this new virus shared around 80∼90% sequence identity to SARS-CoV(2). Both bioinformatics modeling and *in vitro* experiments indicated that 2019-nCoV was more likely to use the angiotensin-converting enzyme 2 (ACE2) as the entry receptor(3, 4). ACE2 is previously known as the receptor for SARS-CoV and HCoV-NL63(5, 6). Considering the sequence of 2019-nCoV is similar to the SRAS-CoV and they are reported to share the same molecular as the entry point, analyses of ACE2 expression and distribution in lung and related biological processes may help us understand the pathogenesis and design therapeutic strategies for 2019-nCoV. Recent advances in bioinformatics enable researchers to reveal the underlying mechanisms of various diseases. In this study, we used bioinformatics approaches to identify the ACE2 expression features in lung and mine associated regulating networks. These results may help us understand the pathogenesis and design therapeutic strategies for 2019-nCoV.

## Results

### ACE2 expression features in lung

Among the 2019-nCoV infected patients, there are many patients with underlying diseases, such as chronic respiratory disease and cardiovascular disease. Therefore, it is important to evaluate whether patients with underlying diseases are more susceptible to coronavirus infection than healthy population. Firstly, we analyzed the expression of ACE2 in lung in the different populations. The expression level of ACE2 was not significantly altered between healthy populations and patients with chronic respiratory diseases including chronic obstructive pulmonary diseases (COPD) and asthma (Figure 1A, 1B, and 1C, p-value > 0.05). It indicated that 2019-nCoV could have the same ability to bind the respiratory tract epithelial cells of healthy population and patients with chronic respiratory diseases through ACE2. Thus, there may be no significant difference in the susceptibility of 2019-nCov infection between healthy population and patients with chronic respiratory diseases.

**Figure 1.**
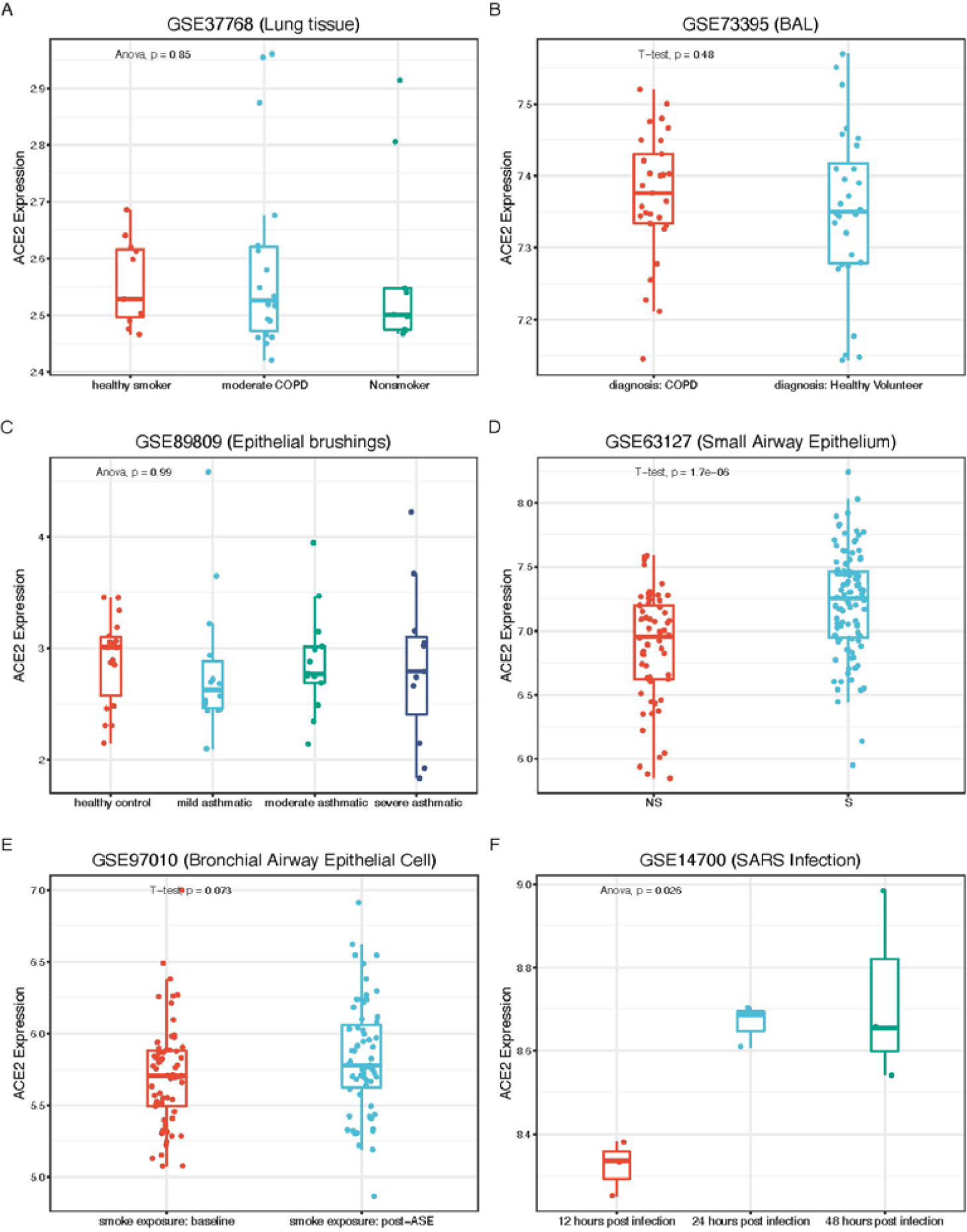
Expression of ACE2 in lung tissues and epithelial cells. A. Expression of ACE2 in lung tissues in healthy smokers, moderate COPD patients and non-smokers. B. Expression of ACE2 in bronchoalveolar lavage samples of COPD patients and healthy volunteers. C. Expression of ACE2 in asthma patients. D. Expression of ACE2 in small airway epithelial in smokers (S) and non-smokers (NS). E. Expression of ACE2 after ASE. F. Expression of ACE2 in SARS-CoV infected bronchial cells.

Secondly, we found that the level of ACE2 expression was markedly upregulated in long-term smokers (Figure 1D, p-value < 0.05). However, the p-value between baseline and post-acute smoke exposure (ASE) group (24 hours after smoking 3 cigarettes) was 0.073 (Figure 1E). It was assumed that the long-term smoking could elevate the expression of ACE2 and thus, increase the susceptibility to 2019-nCoV infection.

Thirdly, we analyzed the expression of ACE2 in airway epithelial cells after being infected with SARS-CoV. The result suggested that 24 hours after SARS-CoV infection, the expression of ACE2 dramatically increased compared to 12 hours. After 48 hours, the expression of ACE2 remained at a high level. This indicated that ACE2 not only played a critical role in viral susceptibility but also may be involved in post-infection regulation.

### Biological function of ACE2 in coronavirus infection

To investigate the virus-related potential biological processes associated with ACE2, we extracted the expression profiles of 15 healthy non-smokers from dataset GSE89809 and divided them into two groups: high and low-expression level of ACE2. We performed GSEA (Gene Set Enrichment Analysis) and found that the expression of ACE2 was mainly associated with innate and acquired immune responses, regulation of B cell mediated immunity, cytokine and IL-1, IL-10, IL-6, IL-8 secretion (Figure 2A). Huang et al. reported that 2019-nCoV infected patients had higher expression level of IL-1B, IL-7, IL-8, IL-10, etc. (1). This implies that the up-regulation of ACE2 could mediate the inflammatory responses which were induced by above-mentioned cytokines. Moreover, higher expression of ACE2 could prolong virus life cycle, enhance virus replication and mediate penetration of the virus into the host cell.

**Figure 2.**
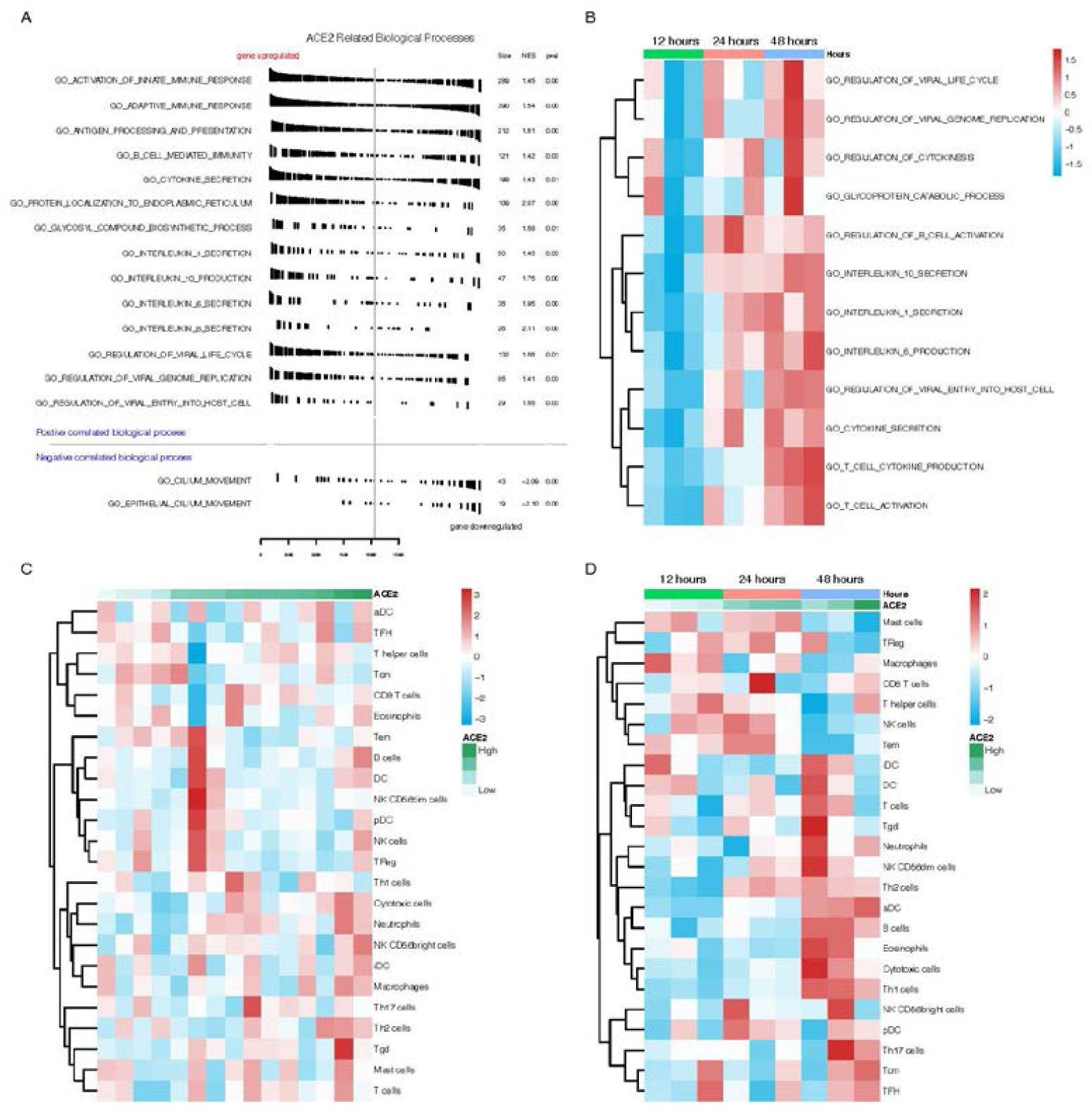
Functional analysis of ACE2 revealing related biological processes. A. GSEA analysis showing the related biological processes of ACE2 in healthy non-smokers. B. GSVA score of key processes showing time-dependent alterations. C. Immune infiltrating in healthy population. D. Immune infiltrating in SARS-CoV infected cells.

To further explore whether above activities could be triggered after coronavirus infection, we analyzed the expression profiles of epithelial cells which was infected by SARS-CoV. The biological process scores were obtained using GSVA (Gene Set Variation Analysis) and the pearson correlation between process scores and ACE2 expression was calculated. Similarly, while ACE2 increased after infection, production of cytokines, IL-1, IL-10 and IL-6 also increased, and the regulation of B cell activation was triggered to response. After 48 hours, virus activities such as viral entry into the host cell, virus life cycle, and viral transcription were enhanced. Besides, the T-cell cytokine secretion was increased, and T-cell activation was promoted (Figure 2B). Then, we used ssGSEA (Single Sample Gene Set Enrichment Analysis) to quantify the immune infiltrates in healthy people epithelial cells and cells after infection. We found that in normal samples, the immune cells were not activated so that the correlation between infiltration and ACE2 expression was not significant (Figure 2C). However, in the SARS-CoV infected cells, the ACE2 was significantly correlated with neutrophils, NK cells, Th17 cells, Th2 cells, Th1 cells, DC (Figure 2D).

### Regulating networks after infection

Considering higher expression of ACE2 was involved in mediating inflammatory responses, immune activities, and viral replication in both healthy individuals and infected cells, we further explored protein-protein regulation network in viral activity-related proteins and cytokine secretion, respectively. We extracted the viral activity-associated and cytokine secretion-associated proteins from the Gene ontology (GO) biological process *gmt* file. Construction of the Protein-protein interaction (PPI) showed that these proteins were tightly interactive among each other and we here exhibited the most significant PPI. In terms of viral activity, we found that *“viral gene expression”* proteins such as RPS3, RPS8 and PRS25 were the most important genes in the PPI network. In addition, VCP, LARP1, UBA52, PRKN, EIF3A and EIF3L were also important in the network (Figure 3A). In terms of the cytokine secretion associated proteins, we found that proteins such as SRC, FN1, MAPK3, LYN, MBP, NLRC4, NLRP1 and PRKCD were Hub proteins in the regulating network of cytokine secretion after coronavirus infection (Figure 3B), indicating that these molecules were critically important in ACE2-induced inflammatory response.

**Figure 3.**
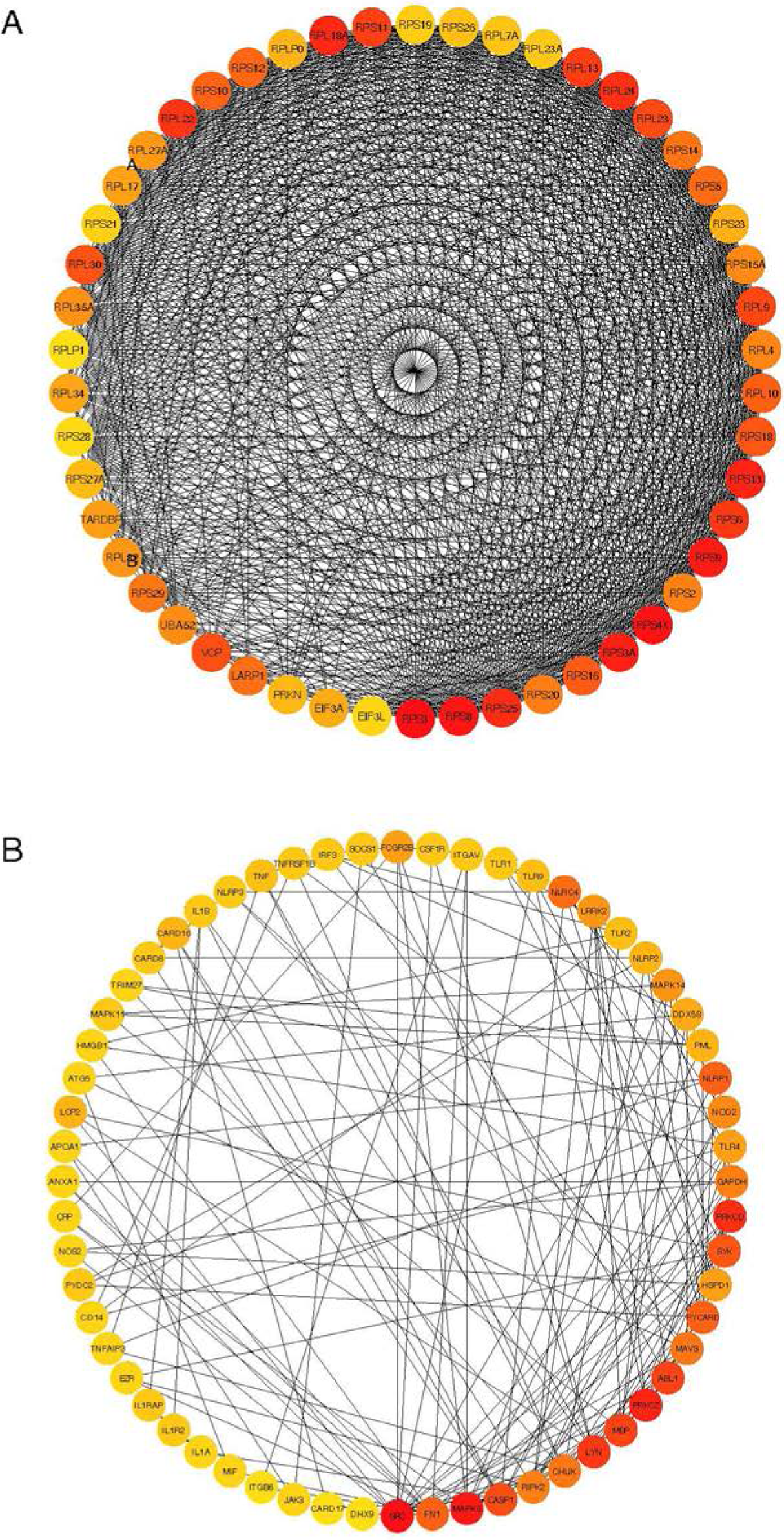
PPI network A: PPI network of viral replication-related proteins. B: PPI network of cytokine secretion-related proteins.

## Discussion

2019-nCoV infection in Wuhan is a serious public health problem. As the number of cases has been increasing and the scope of infection has been expanding, people keep a close eye on the future development of the epidemic. Based on the data of the first 41 patients, the reduction of lymphocytes was a prominent feature of the laboratory examination. The main symptoms of patients with 2019-nCoV infection were interstitial changes in the lung according to the chest CT. Moreover, severe pulmonary interstitial disease led to hypoxemia and type I respiratory distress, which was related to the prognosis of patients. 2019-nCoV invades the lungs through the respiratory tract and causes severe pneumonia, which is the main pathogenesis mode. It was found that 2019-nCoV was able to bind to ACE2 receptor on the surface of epithelial cells. At the beginning of the outbreak, Dong N et al. calculated and simulated that 2019-nCoV may have the same ACE2 receptor as SARS, although its binding ability to ACE2 was weaker than SARS. Zhou P et at. further confirmed that ACE2 was necessary for the cells infected by 2019-nCoV(4). In this study, six independent studies (GSE37758, GSE73395, GSE89809, GSE63127, GSE97010, and GSE14700) were analyzed, covering healthy population, chronic obstructive pulmonary disease patients, bronchial asthma patients and smoking patients. The samples included lung tissue, BALF, airway epithelial cells, small airway epithelial cells and coronavirus infected epithelial cells. We showed that ACE2 was found in all of the samples including healthy population and patients with chronic airway diseases. It revealed that there was no significant difference in the susceptibility of 2019-nCoV between healthy population and patients with chronic airway diseases. The expression of ACE2 was up-regulated in smokers, suggesting that smoking may increase the susceptibility of new coronavirus. Further analysis showed that the expression of ACE2 could be up-regulated by SARS coronavirus infection.

Previous studies have found that ACE2 was related to the severity of acute respiratory syndrome induced by coronavirus infection, which regulated the production of acute respiratory distress syndrome-associated cytokines(7). ACE2 is also related to adaptive immune responses(8). In this current study, the GSEA analysis showed that the high expression of ACE2 was related to innate immune response, acquired immune response, B cell regulatory immunity and cytokine secretion, and enhanced the inflammatory response induced by IL-1, IL-10, IL-6, IL-8 cytokines. We speculated that the immune system dysfunction involved in the high expression of ACE2 was related to the symptoms called cytokine storm. The clinical study in Wuhan pointed that the levels of IL-1B, IL-10 and IL-8 were significantly increased in critically ill patients with new coronavirus infection, indicating that there was cytokine mediated inflammatory responses in patients with coronavirus infection(9). At the same time, our results also showed that the high expression of ACE2 increased the expression of genes involved in viral replication, which was beneficial for the virus to enter the host cells. These gene enrichments were consistent with those of the epithelial cells infected by coronavirus. It suggested that the transcriptome of epithelial cells was altered after the infection of coronavirus though the regulation of ACE2, which was beneficial to the replication and assembly of virus, and also to the entry of virus into host cells. Meanwhile, T cell activation and inflammatory response mediated by T cell factors were significantly induced by the alteration of transcriptome as well.

It was speculated that the increased levels of IL1B, IFN γ, IP10, and MCP1 in patients infected with new coronavirus were linked to the activation of T-helper-1 (Th1) cell responses(9). However, there is a lack of pathological data of lung tissue infected with new coronavirus. Through ssGSEA analysis, we found the correlation between ACE2 and immune cell infiltration. The results showed that the high expression of ACE2 in lung tissue was beneficial to induce cytotoxic reaction, neutrophil inflammation and Th2-dominated immune response. Moreover, the expression of ACE2 was in a time-dependent manner after the virus infection. In the meantime, ACE2 mediated the activations of neutrophils, NK cells, Th17 cells, Th2 cells, Th1 cells, DC cells, TNF - α secreting cells, which induced more serious inflammatory response.

In order to further analyze the core genes involved in viral replication and inflammatory response mediated by ACE2, we constructed a protein-protein interaction network. It was found that RPS3 played a key role in viral replication, and SRC non-receptor protein kinase was the hub gene in inflammatory response. It has been shown that RPS3 plays an important role in cell mitosis by interacting with α - tubulin. When cells are infected with HIV-1, HIV-1 Tat protein can interact with RPS3 to disturb its localization on the spindle, and ultimately inhibit cell proliferation(10). Moreover, the function of RPS3 depends on its own nuclear translocation, which is regulated by protein kinase like SRC(11). We speculated that the increased expression of ACE2 affected RPS3 and SRC, which were the two hub genes involved in viral replication and inflammatory response.

In conclusion, 2019-nCoV infection is a serious public health problem, endangers human health seriously. Due to the strong human to human infectivity, it needs to be further studied. Our current study found that there was no significant difference in ACE2 expression between healthy population and patients with chronic airway disease. The activations of neutrophils, NK cells, Th17 cells, Th2 cells, Th1 cells, DC cells, TNF - α secreting cells were induced by overexpression of ACE2, which participate in the production of more obvious inflammatory response. A schematic model was showed in figure 4. These findings may help clinicians and researchers gain more insights of pathogenesis and design therapeutic strategies for 2019-nCoV. Note all results are based on data mining, thus, further experiments are needed.

**Figure 4.**
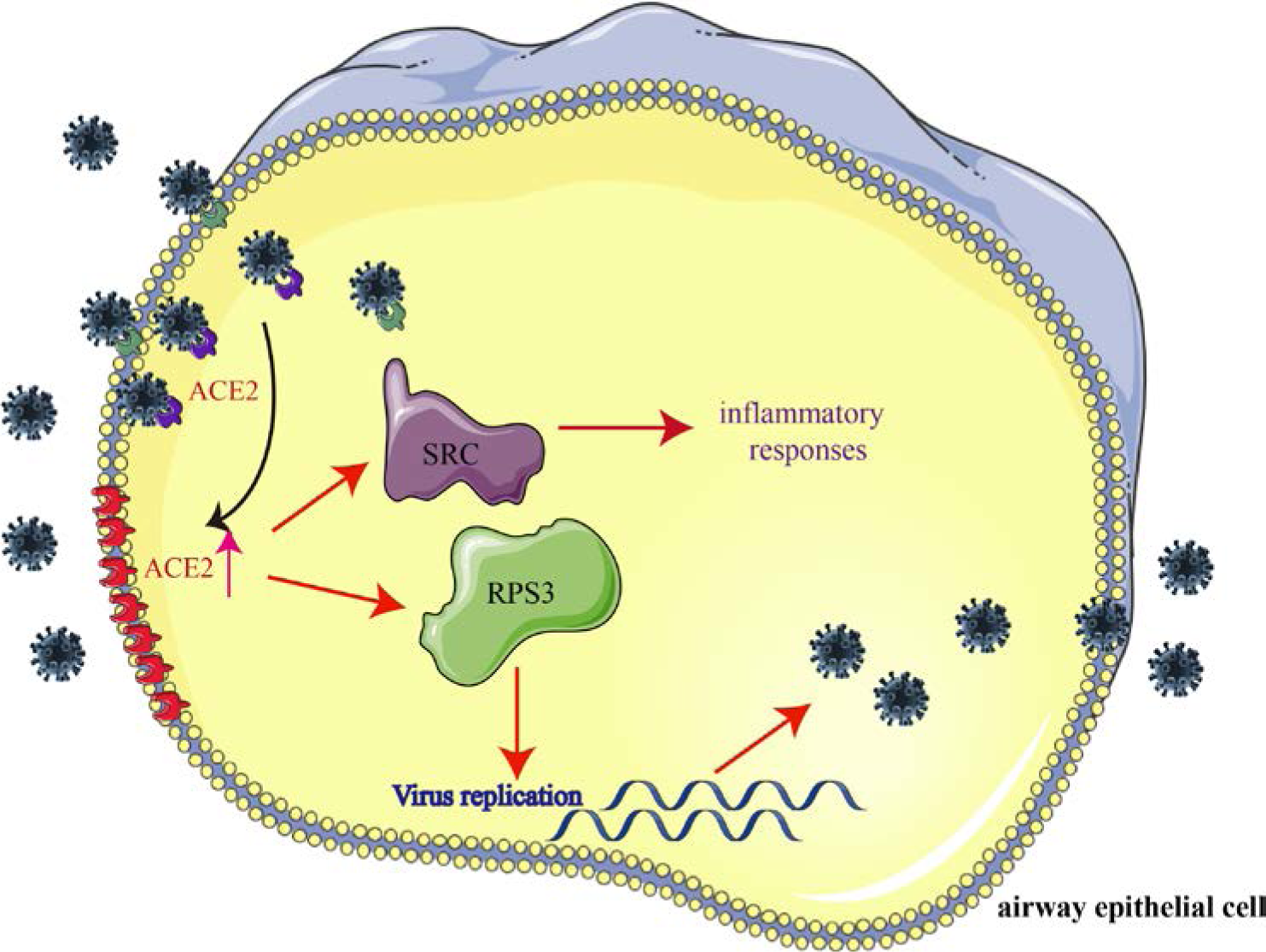
A schematic model of 2019-nCoV infection. 2019-nCoV use ACE2 as a cellular entry receptor in airway epithelial cells. At the same time, the expression of ACE2 is increased by the infection. Furthermore, the increased expression of ACE2 affected RPS3 and SRC, the two hub genes involved in viral replication and inflammatory response.

## Methods

### Data Collection

Six independent studies (GSE37758, GSE73395, GSE89809, GSE63127, GSE97010, and GSE14700), covering healthy volunteers, COPD patients, asthma patients, smoking individuals were collected from GEO data repository (https://www.ncbi.nlm.nih.gov/geo/). The samples consist of lung tissues, bronchoalveolar lavage, bronchial epithelial cells, small airway epithelial cells, and SARS infected cells.

### Functional enrichment analysis

GO biological process annotation *gmt* file was downloaded from MSigDB (https://www.gsea-msigdb.org/gsea/msigdb/). GSEA was performed to analyze the possible biological processes related to ACE2 in healthy people using clusterProfiler (12). The parameters were nPerm = 1000, minGSSize = 10, maxGSSize = 500, the biological processes with p-value < 0.05 were considered significant. GSVA was performed using GSVA R package. The immune infiltrating was quantified using the ssGSEA method in GSVA R package. The gene list for immune cells was derived from Bindea G et al.(13).

### PPI

All viral-related biological process and cytokine secretion-biological process proteins were extracted from the *gmt* file, Cytoscape v3.7.2 was used to construct the PPI network using BisoGenet application, the PPI sources include DIP, BIOGRID, HPRD, INTACT, MINT and BIND. And the nodes with topological importance in the interaction network were screened by calculating Degree Centrality (DC) with the Cytoscape plugin CytoNCA. Hub proteins were identified using Cytoscape plugin CytoHubba.

## Data Availability

yes

## Acknowledgements

The work was supported by 2019-nCoV tackling project of Chengdu Science and Technology Bureau.

